# Genetic risk factors for death with SARS-CoV-2 from the UK Biobank

**DOI:** 10.1101/2020.07.01.20144592

**Authors:** Chang Lu, Rihab Gam, Arun Prasad Pandurangan, Julian Gough

## Abstract

We present here genetic risk factors for survivability from infection by the severe acute respiratory syndrome coronavirus 2 (SARS-CoV-2) responsible for coronavirus disease 19 (COVID-19). At the time of writing it is too early to determine comprehensively and without doubt all risk factors, but there is an urgency due to the global pandemic crisis that merits this early analysis. We have nonetheless discovered 5 novel risk variants in 4 genes, discovered by examining 193 deaths from 1,412 confirmed infections in a group of 5,871 UK Biobank participants tested for the virus. We also examine the distribution of these genetic variants across broad ethnic groups and compare it to data from the UK Office of National Statistics for increased risk of death from SARS-CoV-2. We confidently identify the gene ERAP2 with a high-risk variant, as well as three other genes of potential interest. Although mostly rare, a common theme of genetic risk factors affecting survival might be the inability to launch or modulate an effective immune and stress response to infection from the SARS-CoV-2 virus.

## Introduction

At the time of writing the world is gripped in global pandemic of such proportions that the economic cost is being compared to the second world war. It is abundantly clear that the main risk factors for survival from the infection by SARS-CoV-2 are age and pre-existing conditions. There may still be important genetic risk factors however, and determining these could contribute by identifying vulnerable people, be relevant to vaccination^1^, suggest drug targets, and gain a deeper understanding of the biology determining survival in patients. Furthermore, there is an important social question raised by data from the UK Office of National Statistics (ONS) ^2^ showing that some ethnic groups, black (black Caribbean, black African, black Other), had an elevated risk of death from SARS-CoV-2 infection relative to European and South Asian ethnic groups. Any bias in ethnic distribution of risk factors will contribute to the discussion on the balance between social or genetic factors driving the measured ethnic imbalance in survival rates.

Once people become infected with the coronavirus the following can happen: they do not manifest any symptoms (asymptomatic), either recover with mild or severe complications (e.g. respiratory failure) requiring hospitalisation, or they succumb. There are several studies reporting COVID-19 genetic research ^3,4^ addressing susceptibility to infection in the population, and only one recent study goes further and looks at the second possible outcome by examining those who suffer respiratory failure ^5^. We chose to examine the third possible outcome and determine genetic risk factors for death with SARS-CoV-2.

Within half a year, the first dedicated large-scale sequencing studies of the COVID-19 outbreak will be available, and within a full year there will be enough genetic data to comprehensively assess risk factors. But even now at the time of writing we are only 4 months from the pandemic being declared by the World Health Organisation. The only way to advance genetic research more quickly in this moment of urgency is to use cohorts for whom genetic data already exists. National biobanks such as the Spain and Italy biobanks used in one recent study ^5^, have access to the most reliable clinical data and outcomes, whereas commercial providers such as 23andMe who have done their own study have greater numbers ^6^ but lack accurate clinical data.

We present here our analysis focused on protein-coding gene variants in the first release of UK Biobank data including clinical outcome as well as infection test results. It is challenging to find significant causal variants from the group of 5,971 participants tested for COVID-19, which include 193 deaths from 1,412 people testing positive. In order to attain early results, we have leveraged information on functional ontologies for genes and on deleteriousness of substitutions on top of association statistics. Our study departs from the traditional Genome Wide Association Study (GWAS) because correlation alone is insufficient to find significant associations.

## Results

We have discovered 4 genes (Table 1) with missense variants that are significant, and one of those genes also contains an additional non-coding variant in an intronic region, which may not be significant in isolation but worth noting in the context of the independently identified gene.

**Table 1.**
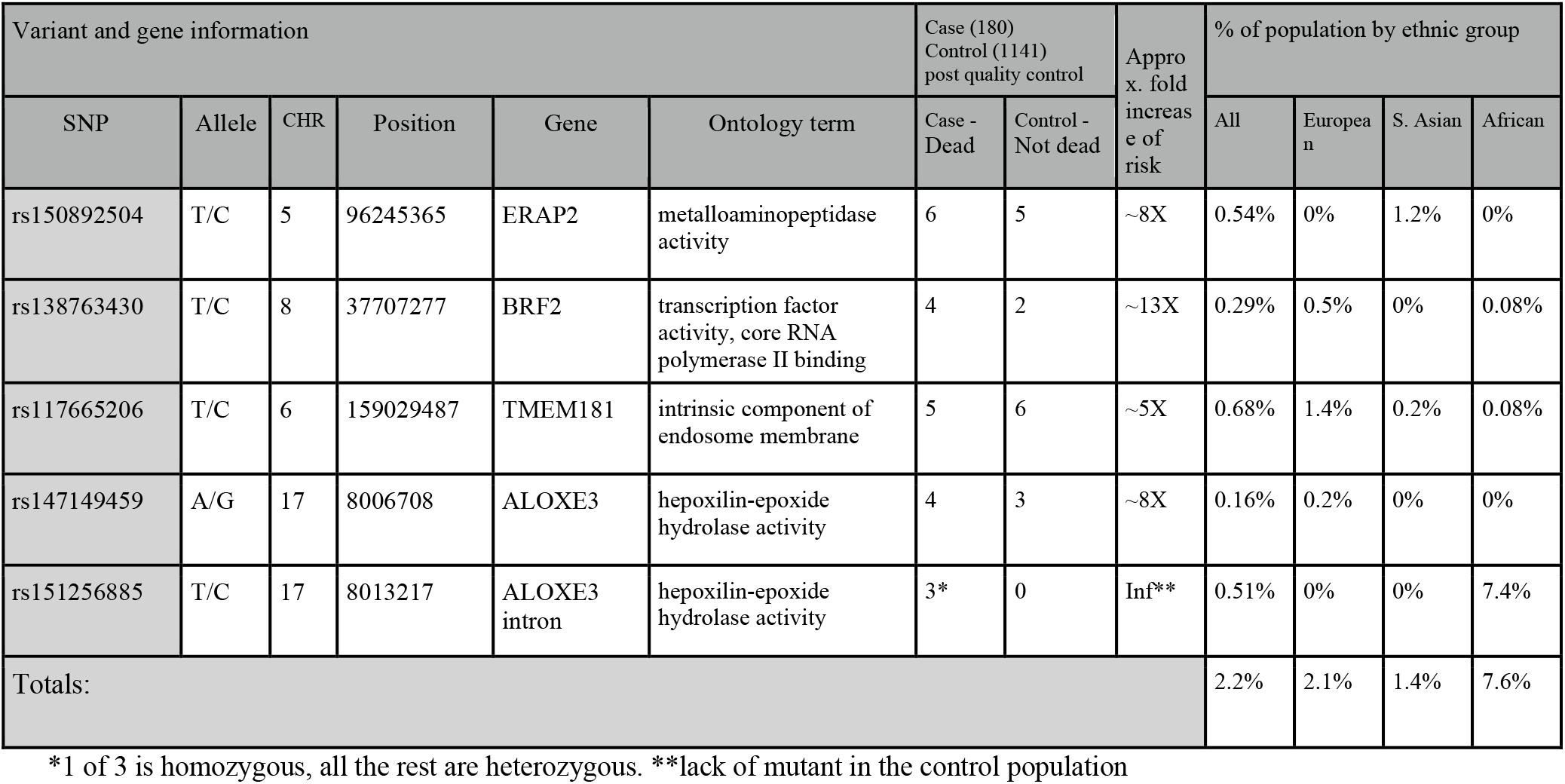
Genetic risk variants. This table shows the 5 genetic variants we have identified as potentially significant risk factors for death from SARS-CoV-2 infection. Coordinates are from the GRCh37 assembly of the human genome. Ontology terms are those predicted by dcGO, and case/controls are after the quality control was applied (see methods). Allele frequencies in different ethnic groups were taken from the 1000 genomes project, and the gross frequencies in the ‘All’ column taken from ALFA (https://www.ncbi.nlm.nih.gov/snp/docs/gsr/alfa/) for greater accuracy.

| Variant and gene information |  |  |  |  |  | Case (180)<br>Control (1141)<br>post quality control |  | Approx. fold increase of risk | % of population by ethnic group |  |  |  |
| --- | --- | --- | --- | --- | --- | --- | --- | --- | --- | --- | --- | --- |
| SNP | Allele | CHR | Position | Gene | Ontology term | Case - Dead | Control - Not dead |  | All | European | S. Asian | African |
| rs150892504 | T/C | 5 | 96245365 | ERAP2 | metalloaminopeptidase activity | 6 | 5 | ~8X | 0.54% | 0% | 1.2% | 0% |
| rs138763430 | T/C | 8 | 37707277 | BRF2 | transcription factor activity, core RNA polymerase II binding | 4 | 2 | ~13X | 0.29% | 0.5% | 0% | 0.08% |
| rs117665206 | T/C | 6 | 159029487 | TMEM181 | intrinsic component of endosome membrane | 5 | 6 | ~5X | 0.68% | 1.4% | 0.2% | 0.08% |
| rs147149459 | A/G | 17 | 8006708 | ALOXE3 | hepoxilin-epoxide hydrolase activity | 4 | 3 | ~8X | 0.16% | 0.2% | 0% | 0% |
| rs151256885 | T/C | 17 | 8013217 | ALOXE3 intron | hepoxilin-epoxide hydrolase activity | 3* | 0 | Inf** | 0.51% | 0% | 0% | 7.4% |
| Totals: |  |  |  |  |  |  |  |  | 2.2% | 2.1% | 1.4% | 7.6% |

\*1 of 3 is homozygous, all the rest are heterozygous. \*\*lack of mutant in the control population

Overall, we can see that none of the missense variants are very common in the general population, and we also observe that some of the variants are clearly over-represented in one of the broad ethnic groups defined in the ONS data. Taken together these high-risk variants potentially cover 2.2% of the general population as represented by ALFA (aggregate allele frequency from dbGaP (https://www.ncbi.nlm.nih.gov/gap/). We observe that non-coding intronic variant rs151256885 is seen almost exclusively in people of African ethnicity, the highest risk group, and that it is quite common in that group (7.6%). With the amount of data we have it is premature to say that there is an ethnic bias in genetic risk.

### ERAP2

The endoplasmic reticulum (ER) aminopeptidase 2 (ERAP2) codes for a zinc metalloaminopeptidase, a member of the oxytocinase M1 ^7^. This gene plays a major role in N-terminal trimming antigenic epitopes for the making of HLA class I binding peptides to be presented by the major histocompatibility complex class I (MHC-I) molecules ^8^. This trimming step is essential to tailor longer precursor peptides bringing them to the correct length needed for their presentation by the MHC-I molecule ^9^. All infectious processes occurring in the intracellular space are strictly processed and presented by these MHC-I molecules, thus alerting the immune system and providing targets for cytotoxic T cell response. Consequently, vaccine designs and optimisation based on the MHC-I presented peptide epitopes could induce the immune responses bettering the survival chances ^10^. The gene is expressed broadly across most cell types, but with a reduced level of expression only in the brain ^11^.

ERAP2 has been investigated in the case of infection with SARS-CoV-2 alongside two other enzymes, ERAP1 and IRAP, and has been demonstrated as the most stable of the enzymes generating 8-10 residues-long antigenic peptides which is the optimal length for HLA binding. The effectiveness of this process reflects effectiveness of the immune response to be generated upon infection with SARS-CoV-2; an aberrant generation of antigenic peptides can lead to the infection evading the immune system ^12^.

ERAP2 along with ERAP1 are highly polymorphic genes with various naturally occurring variants that have been shown to play major roles in several pathological conditions from bacterial and viral infections to cancer ^13,14^. The mutation R751C is disruptive to the fold of the protein (Figure 1A). The wild type arginine is solvent inaccessible and buried to 5.3 Å in a well-packed environment with occluded surface packing (OSP) density of 0.5, making 17 contacts within a 10Å radius. Residue depth and packing density measures have been shown to be an important parameter to study mutation and its impact on structures ^15^. The mutant cysteine does not fill the cavity, only engaging in 5 contacts, thus destabilising the protein. This could be a potentially druggable target for treatment with Cysteamine^16^ in patients with this variant who contract the disease.

**Figure 1.**
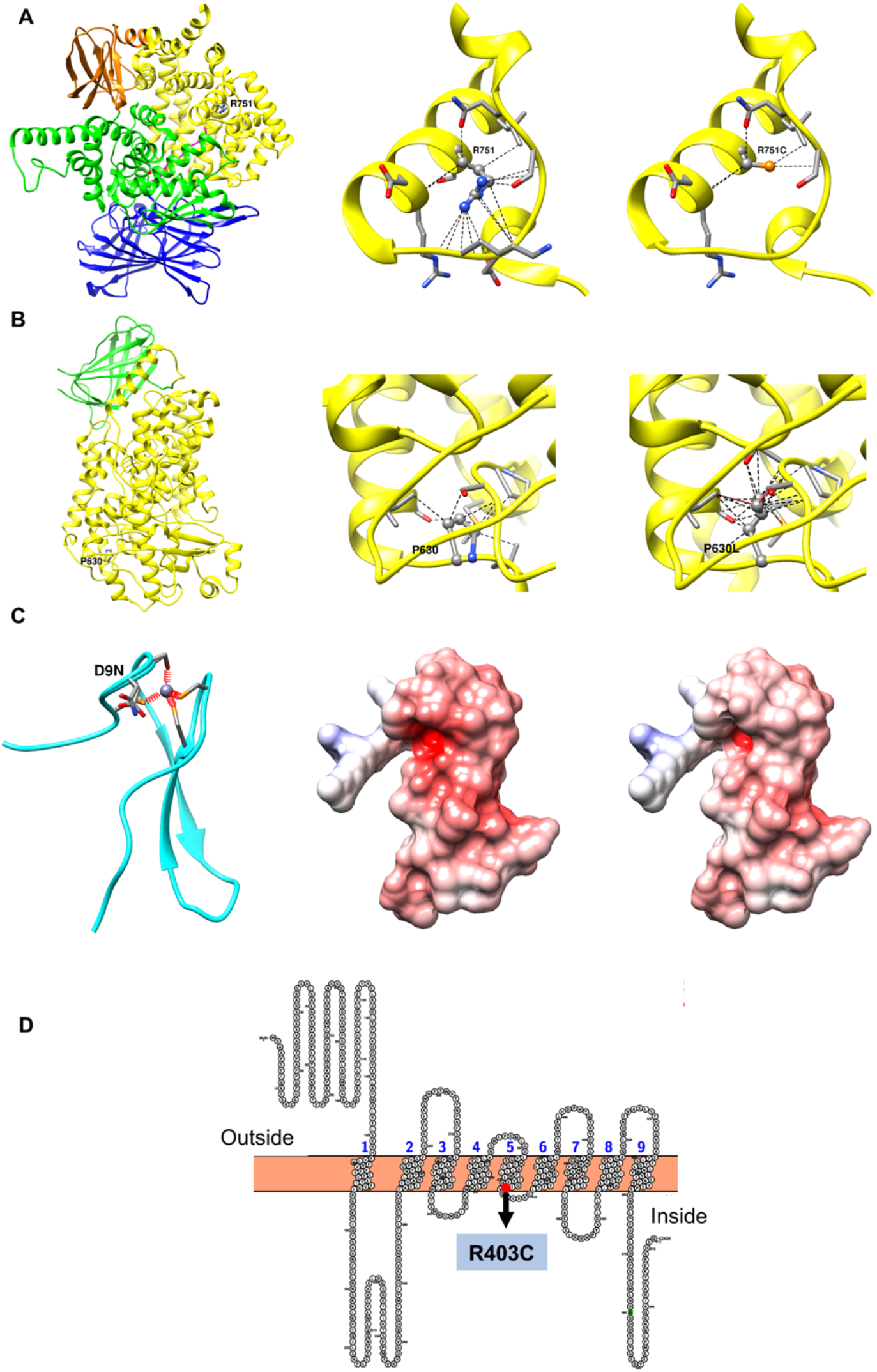
Structural analysis of mutations. (A - left) The wildtype residue R751 of gene ERAP2 mapped onto PDB structure ^41^ (PDB ID 5AB0). The domains 1, 2, 3 and 4 are coloured blue, green, orange and yellow respectively. (A – centre) shows the number of contacts formed by the side chain atoms of R751 with the neighbouring atoms within 10 Å radius. (A – right) shows the number of contacts formed by the side chain atoms of the mutant R751C with the neighbouring atoms within 10 Å radius. (B - left) The wildtype residue P630 of gene ERAP2 mapped onto the homology model based on PDB 4NRE). (B – centre) shows the number of contacts using Chimera ^42^ formed by the side chain atoms of P630 with the neighbouring atoms within 10 Å radius. (B – right) shows the number of contacts formed by the side chain atoms of the mutant P630L with the neighbouring atoms within 10 Å radius. Wildtype and mutant residues are shown in ball and stick representation. All other residues in contact with wildtype and mutant residues are shown as stick representation. (C - left) Zn Ribbon domain of BRF2 (aa 3-40) showing the mutation D9N along with the zinc ion coordinated by 4 cysteine residues in orange. (C – centre) Electrostatic potential surface areas of the wildtype Zn Ribbon binding domain. (C – right) Electrostatic potential surface areas of the mutant Zn Ribbon binding domain. Electrostatic potential was calculated using Adaptive Poisson Boltzmann Solver ^43^ (APBS) and rendered using Chimera ^42^. (D) Transmembrane topology for TMEM181 showing 9 transmembrane helices predicted by TMHMM and depicted using the program ^44^. The mutant R403C is found on the 5^th^ transmembrane helix close to the intra-cellular membrane region (annotated and highlighted as red dot).

### BRF2

The Transcription factor IIIB 50 kDa subunit (TFIIIB50/BRF2) gene is an activator of the RNA polymerase III (Pol III) transcription and a member of the TFIIB family of transcription factors (TFs), and has been identified as a lineage-specific oncogene in lung cancer ^16^. The TFIIB-related factor 2 (BRF2) is involved in the essential gene transcription process recruiting the Pol III to type III gene promotors ^17^. This requires an accurate recruitment of this polymerase alongside the target genes thus forming the pre-initiation complex (PIC). TFIIIB has been demonstrated to be a key factor for this process ^18-20^. Tight regulation is imposed on the of BRF2- Pol III mechanism during the different phases of the cell cycle with upregulation of BRF2 is linked to very low survival rates in patients with various types of carcinomas with studies showing that BRF2 is a redox-sensing factor playing a major role in the response to oxidative stress promoting cancer cell evasion of the immune system under this condition ^21-23^. Oxidative stress or redox imbalance, a phenomenon that is considerably increased with aging, has been previously linked to respiratory viral infections ^24^ and is a key player in Sars-Cov-2 infection. Abundant production of reactive oxygen species (ROS) along with the degradation of the antioxidant mechanism makes a rich ground for viral replication inclusive of Sars-Cov-2 ^25^. BRF2 is broadly expressed across all tissues in GTEx ^11^ confirming its generalised role in stress response.

The protein has a zinc finger domain at the N terminus, the cyclin-like transcription factor IIB domain in the core, and a short disordered C-terminal region with phosphorylation sites and recognition motifs. Structural analysis shows that the mutation D9N at the Zn Ribbon domain considerably alters the electrostatic potential surface which in turn directly impacts the fundamental property of the domain to recognise nucleotide binding partners (Figure 1C). Thus this variant most likely negatively alters the selectivity of the protein.

### ALOXE3

The arachidonate lipoxygenase 3 (ALOXE3), located on chromosome 17p13.1, and is a member of the lipoxygenase family (LOXs) ^26,27^. This family of lipid peroxidation enzymes has been implicated in key cellular processes such as proliferation and differentiation, and also in infectious diseases with pro-inflammatory effects ^27-29^. ALOXE3 encodes for two LOX isoforms both expressed in the skin and is proposed to play a major role in the development of the skin permeability barrier ^28,30,31^. This could affect susceptibility to infection but is no explanation for a high death rate post infection.

Although normally expressed in the skin ^11^, under viral infection the GEO signatures database for differentially expressed genes shows ALOXE3 is slightly up-regulated under ebolavirus infection and even more up-regulated by SARS-CoV ^32^ in human airway epithelial cultures. Thus, although there is no definitive explanation for the mechanism for a role in SARS-CoV- 2 survivability within our current knowledge of molecular biology, our genetic study combined with the differential expression observed in SARS-CoV, may be enough evidence to suggest an undiscovered role for this gene. When compared with other LOX isoforms involved in fatty acid metabolism, ALOXE3 has limited oxygenase activity, so it has been suggested to function as a fatty acid peroxide isomerase. One hypothesis is that ALOXE3 may be co-opted by viruses to render the specific lipid microenvironment adequate for multiplication. The LOX family are reported as inflammatory mediators, so this gene could also be modulating immune response and play a part in failure to suppress immune response leading to the cytokine storm that is frequently reported in SARS-CoV-2 patients.

The wildtype residue P630 of our variant is completely buried at the residue depth of 11.3 Å and has an OSP value of 0.53. It makes a total of 10 contacts (Figure 1B – top right). Mutant P630L represents a bulky replacement in an already well packed environment. Due to the substitution of larger amino acid Leucine, the number of contacts increases to 22 in the structural model at the expense of two serious steric clashes with the neighbouring residues S608 and I634 (shown as red lines in Figure 1B – bottom right). Bulky mutations in a buried and well packed environment are usually not tolerated and lead to protein destabilisation.

In addition to the significant disruptive missense variant in this gene, there is also an intronic noncoding variant. Although this variant was not significant at the 95% confidence level in the GWAS calculation, it was one of only 6 variants in the entire genome above the baseline (significant at the 50% level, suggesting half may be true). Since it falls in the same gene as one of the 4 significant missense variants, it is unlikely to be coincidence and worthy of mention in the results here. The variant is only 16 bases downstream of the exon splice junction and could cause cryptic splicing. The variant is predicted to be damaging with a probability of 0.61 by regSNP-intron ^33^ and classified on the borderline between ‘damaging’ and ‘possibly damaging’, which is consistent with the possibility of this variant having a functional effect.

### TMEM181

The Transmembrane Protein 181 (TMEM181, also known as GPR178, KIAA1423), located on chromosome 6q25.3, encodes a protein expressed on the cell surface ^9^. The mutant R403C is predicted to be found on the 5th transmembrane helix close to the intra-cellular membrane region (Figure 1C). An analysis of disease mutations in human transmembrane proteins suggest that frequent mutations to positively charged amino acids like Arginine have detrimental impact on structure and function of the protein ^34^. Buried charged residues found at or close to the ends of the helices, such as R403C, are involved in catalysis, ligand binding or in stabilising helix caps. There is not much information on this protein in the literature and little to link it to SARS-CoV-2 survivability. What is known about this gene is that it mediates action of cytolethal distending toxins (CDT) which are released by some specific pathogenic bacteria ^35^. TMEM181 is broadly expressed in most tissues (GTEx ^11^), consistent with an innate immune role, however we also observe an additionally elevated expression in the tibial artery; there have been multiple reports ^36-38^ of unexplained lower limb ischemia in COVID-19 patients. We also observed that all 5 of the deaths in people with this variant were in men who self-reported as having hypertension; of the 6 survivors 4 are female and 2 male (one with hypertension). The Human Protein Atlas has it localised within the cell to the vesicles and also Golgi apparatus, the pH of which is altered by coronavirus to protect the spike protein ^39^. As with ERAP2 the Arginine to Cysteine mutation makes this another potentially druggable target with Cysteamine ^40^ for patients with this variant.

## Methods

The SARS-CoV-2 test results and death data from UK Biobank were released on 12 Jun 2020 ^45^. Out of 5871 people tested for the virus who had already been genotyped using the UK Biobank Axiom Array containing 820,967 genetic variants, 1412 were tested positive at least once. These were subjected to data quality Control using Plink software ^46^. We excluded any person missing more than 20% of variants and any variants missing more than 20% of the people (18402 variants and 27 people). We removed any person whose sex in the database had a discrepancy with the genetic data (none found), and anyone with a Hardy-Weinberg equilibrium p value <1e−10 in cases and <1e−6 in controls (13348 variants). We excluded individuals with a high or low heterozygosity rate (195 people) and removed 5 people who were related (pi-hat threshold of > 0.2 in Plink). At the end of QC, there remained 773,676 variants and 5644 people tested for the virus, where 1324 people tested positive, of which 183 have died. We further excluded 3 people who have tested positive with SARS-CoV-2 and have died, but with death-cause possibly unrelated to SARS-CoV-2 (Supplementary Table 1). Finally, the data set contains 180 cases and 1141 controls.

We ran a standard GWAS (Figure 2A) and applied the Holm-Bonferroni multiple hypothesis correction (Figure 2B) that showed no significant associations, but it did show 6 variants that were more significant than all the others (see supplementary data). To overcome this we applied our research group’s experience in protein-coding and hidden Markov models (HMMs) to do a more focussed analysis and go beyond mere statistical correlation by leveraging independent information on gene ontology (dcGO) ^47^ and amino acid substitution deleteriousness using FATHMM ^48^. We used a method of phenotype prediction ^49^ which combines ontologies from dcGO and the same principles of HMM dirichlet mixtures as used by FATHMM, against a genetic background from the 1000 genomes project. From the predicted phenotypes in the UK Biobank cohort participants who were tested for SARS-CoV- 2, potentially causal genes were obtained from the ontology via the dcGO mapping, comprising of: ALOXE3, BRF2, ERAP2, MPP5, RAMP3, RBL1 and TMEM181. Independently of this, we used a separate list of potentially causal genes that was created by the UniProt ^50^ database team, and annotated by them as SARS-CoV-2 receptors comprising: ACE2, BSG, BST2, FURIN, IL6, IL6R, IL6ST, ITGAL, KPNA2, MPP5, PHB2, SGTA, SMAD3 and TMPRSS2 (https://covid-19.uniprot.org/uniprotkb?query=*).

**Figure 2.**
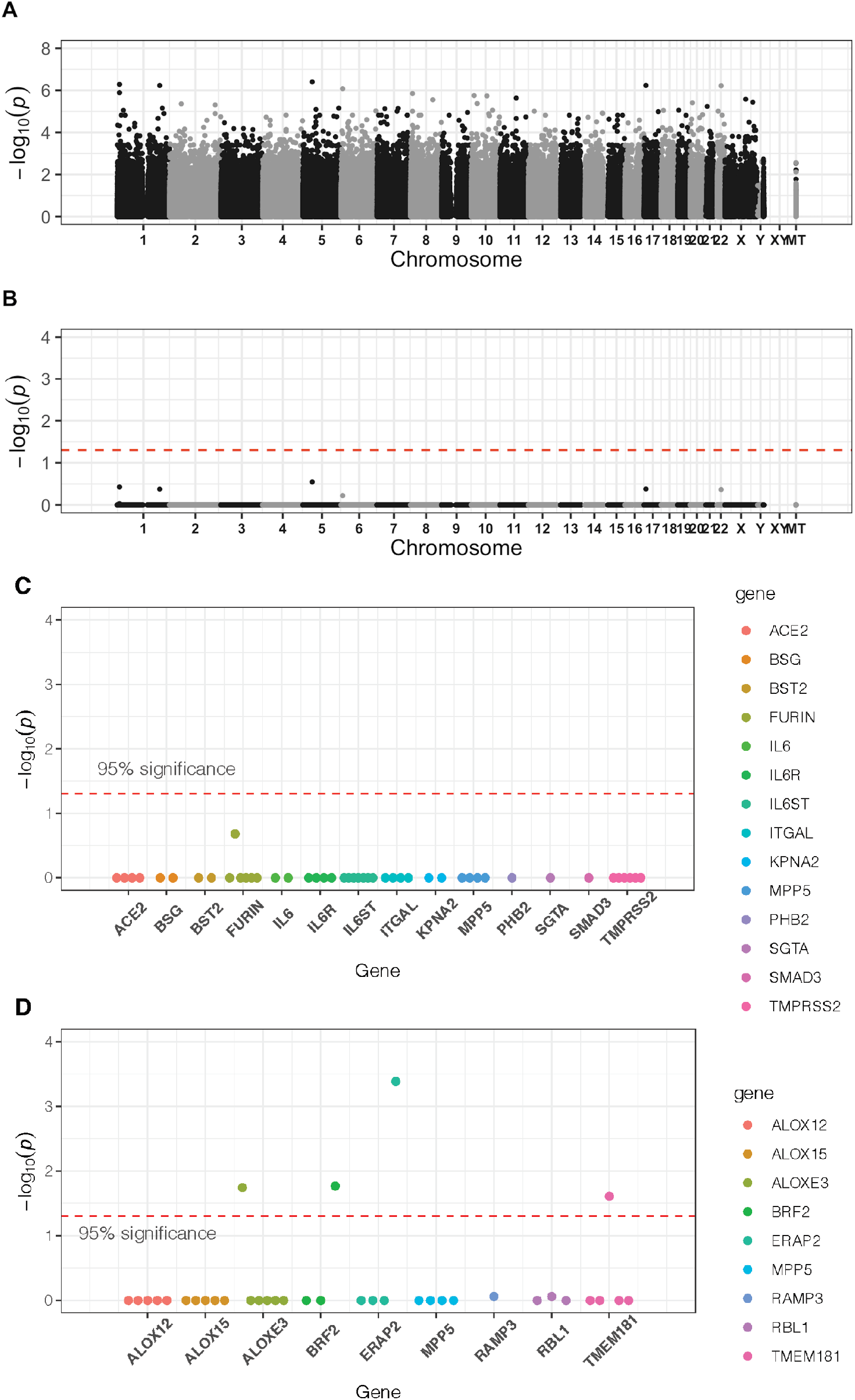
Calculation of variant statistics. (A-B) GWAS summary Manhattan plot of the association statistics for death prognosis after SARS-CoV-2 infection. The association P-values are shown in (A), multi-hypothesis corrected P-values with Holm-Bonferroni method are shown in (B). The red dashed line indicates the genome wide significance threshold of corrected P-value of 0.05. (C) 14 genes known to be associated with Covid-19 selected from literature by the UniProt dataset with a multi-hypothesis corrected, Fisher-combined association and hidden Markov model deleteriousness P-value for each exon variant in the UK Biobank axiom array inside these genes. No variant investigated was found to be significant. (D) Similar combined P-value for exon variants in our selected gene list from ontology-based gene prioritisation using dcGO ^51^.

For both of the two gene lists we applied the following procedure (Figure 2C and Figure 2D). To give additional power to the analysis, we combined the P-value of association from the GWAS calculation (Figure 1A), with the P-value from the HMMs for the deleteriousness of the amino acid substitution. This HMM P-value is taken from the internal probabilities of the two amino acids (mutant and wild) in the HMM for the gene in question ^48^, to obtain a probability of the substitution being tolerated in nature. To combine these two probabilities, we used Fisher’s method for combining P-values. We corrected for multiple hypothesis testing using the Holm-Bonferroni correction, and identified 4 variants as significant at the 95% level of confidence in our gene list (Figure 1C and table 1). The UniProt gene list did not yield any variants significant at the 95% level, but did show one variant that was elevated in significance relative to the others in the FURIN gene.

## Discussion

It is a dangerous time for scientists to present early results on small amounts of data, especially when there is certainty that within a year there will be a lot more data, that will overwhelmingly prove or disprove early findings. However due to the urgency of the global situation, as scientists we have a duty to present any new findings in a timely manner to the best of our knowledge on the available data at the time.

We have identified 4 missense and 1 intronic non-coding variant in 4 genes that significantly increase risk of death in patients contracting SARS-CoV-2. These variants are not common, but together could be present in 2.2% of the UK population (Table 1). Furthermore this information may be of interest to those working on vaccines. We also identified several other variants, missense and non-coding, and other genes which were not significant which merit being highlighted for further investigation when more data is available (supplementary material). In particular aminopeptidase proteins came up several times as potentially being implicated in risk from infection. We also found that variants from two of the genes, those found in ERAP2 and TMEM181, are an Arginine to Cysteine mutation with druggable potential by Cysteamine ^40^.

We do not present any analysis of susceptibility to infection because we would not have a control group. We do not know who has been exposed to the virus. We do not believe that there is enough data to confirm the hypothesis of a genetic explanation to the imbalance in survival rates in the UK as published by the ONS, despite our genetic risk variants being similarly distributed over ethnic groups. The non-coding variant in the intronic region of ALOXE3, common to 7.6% in the African population is definitely worth checking carefully on a larger dataset when it becomes available, since all 3 people with the variant died.

What we have discovered are mostly rare variants, and it is likely that as more data emerges that some more will be discovered. In comparison to other larger statistical association studies, our more ontology-targeted work focused on proteins has yielded rare but high risk variants from several pathways collectively involved in immune and stress response to SARS-CoV-2 infection. Although the infection is a constant across cases, the patients’ individual genetic risk factors can be various. The genes in which we confirmed genetic risk variants provide insights into new biological pathways that are relevant for understanding the severity of the phenotype. Although the sample sizes are small, the risk variants nonetheless have utility in identifying people who may be vulnerable without being aware of it, and who would otherwise go unnoticed.

It is our hope that publishing these tentative discoveries of risk variants well in advance of the inevitable full-scale studies, based on the data we have now, will make a worthwhile contribution to the advance of SARS-CoV-2 infection research and medical advancement.

## Data Availability

The data is from the UK Biobank and is available through them.

## Acknowledgements

We thank Prof. Nic Timpson, Dr. Matt Oates and Dr. Madan Babu for comments on the manuscript. This research has been conducted using the UK Biobank Resource *under Application Number ‘45329’*.

